# Spatiotemporal epidemiology of HIV between 1997 and 2020 in Kerman, Iran

**DOI:** 10.1101/2021.10.21.21265334

**Authors:** Hossein Mirzaei, Ali Mirzazadeh, Mohsen Barouni, Ebrahim Ranjbar, Sana Eybpoosh, Hamid Sharifi

## Abstract

**Objectives:** We assessed the epidemiological and spatiotemporal patterns of HIV cases reported between 1997 and 2020 in Kerman, Iran.

**Method:** We compared the age, gender, modes of transmission, and spatial distribution of newly diagnosed HIV-infected people over time (1997-2004, 2005-2012, and 2013-2020). We also mapped the services and spatial distribution for people who were living with HIV in 2020. The nearest neighbor index and kernel density were used to identify the spatial distribution of HIV cases. Chi-square for trend, one-sample t-test, and Kruskal–Wallis H tests were used to compare the differences over time. Stata 17 and ArcGIS 10.3 were used for HIV surveillance data analysis.

**Results:** A total of 459 (25.6% women) people were diagnosed with HIV during 1997-2020. The proportion of women (from 9.3% in 1997-2004 to 48.3% in 2013-2020, *P* < 0.001), those infected with HIV through sexual contacts (from 11.6% in 1997-2004 to 50.3% in 2013-2020, *P* < 0.001), proportion of HIV infected children under 5 years of age (0.8% in 1997-2004 to 5.4% in 2013-2020, *P* = 0.01), and mean age at diagnosis among men (from 34.9 in 1997-2004 to 39.8 years in 2013-2020, *P* = 0.004) significantly increased over time. The mean age at diagnosis among women (34.5 years in 2013-2020), and the proportion who diagnosed with CD4 count <200 (36.2% in 2013-2020) did not change significantly over time. The density map of residents with HIV infection had the highest density in the northern and southern edges of the city in 2005-2012. The HIV clusters in 2020 matched with the locations of fixed and mobile services.

**Conclusion:** We observed important changes in HIV epidemiology regarding gender, modes of transmission, number of pediatric cases, and density maps over time which should be considered for precise targeting of HIV prevention, treatment, and outreach programs in this city.

Key Messages

- The proportion of women, those infected with HIV through sexual contacts, pediatric cases, and the mean age at diagnosis among men significantly increased over time.
- The density map of residents living with HIV had the highest density in the northern and southern edges of the city in 2005-2012.
- The HIV clusters in 2020 matched with locations of fixed and mobile services.

## INTRODUCTION

HIV epidemic in Iran is concentrated among key populations including people who inject drug (PWID) (15.2% in 2010, 10.9 in 2014, and 3.6% in 2020)^1-3^ and partners of PWID (2.8% in 2010), ^4^ female sex workers (FSW) (2.1% in 2015),^5^ and prisoner (1.2% in 2014), ^6^ but the prevalence of HIV is very low in the general population (0.15%),^7^ blood donors (3 per 100,000 donations from 2003 to 2017) ^8^ and pregnant women (0.04%). ^9^

The Joint United Nations Programme on HIV/AIDS (UNAIDS) estimated 59,314 people living with HIV (PLWH) in Iran in 2019, of whom only 22,054 (37%) were diagnosed and reported to the National HIV Case-Based Surveillance. ^10^ Among the diagnosed cases, 67% (25% of estimated cases) were under antiretroviral treatment, and 85% of treatment cases (11% of estimated cases) had viral load suppression. The gap diagnosis of PLWH is the main gap in the national HIV program in Iran. ^10^ Among patients who were diagnosed with HIV in Iran from 1967 to 2018, most of them were male (83%), aged between 16 and 40 years old (67.6%), and infected through injecting drug use (61.9%). ^9^

Our recent analysis of the HIV continuum of care in Kerman showed that of 1113 estimated number of PLWH in 2016, 437 (39.3%) were diagnosed, 10.1% were on antiretroviral treatment, and 7.3% had viral suppression. ^11^ Beside this cascade analysis, the epidemiological patterns, spatial and temporal changes in HIV epidemiology have not been studied yet; This information is critical for precise targeting of HIV programs in Kerman. In this paper, we used HIV surveillance data from all people diagnosed with HIV in Kerman between 1997 and 2020 to assess the trend in the HIV epidemiological and spatial patterns.

## METHODS

### Study design and data

We used cased-based surveillance data of all HIV cases reported in Kerman city between 1997-2020. Demographic data, age, gender, residential address, first CD4 cell count at the time of diagnosis, modes of transmission (MOT), and date of diagnosis were extracted from the HIV surveillance database. To assess the trend, we analyzed the spatial and epidemiolocal data by three equal time intervals of 1997-2004, 2005-2012, and 2013-2020.

### Study location and environment

Kerman city is located in the southeast of Iran, with a population of around 540,000 individuals. ^12^ One voluntary HIV counseling and testing (VCT) center is responsible for providing care to all people diagnosed with HIV in Kerman. The VCT is the referral center for all people who test positive (or reactive) for HIV at any health facility, DIC (Drop-in center), blood transmission organization, or laboratory in Kerman. For any new referral case, the VCT center collects and sends blood sample for confirmatory HIV diagnosis to a central public health laboratory in Kerman. After confirmation, the VCT staff will submit data to the electronic portal of the National HIV Surveillance. Kerman medical university is supervising the VCT and central public health laboratory.

### Spatial data analysis

We used Google earth for Georeferencing the residential addresses of HIV-infected people (converting a written address to a position on the map surface). ^13^ The nearest neighbor index (NNI) was used to identify if the distribution of diagnosed HIV cases was either random or clustered. The nearest neighbor index was calculated as the observed average distance divided by the expected average distance. ^14^ We produced density maps over the study periods by Kernel density method. ^15^ The ArcGIS version 10.3 was used for spatial data analysis.

### Epidemiological data analysis

For continuous variables (e.g., age, CD4 count), mean, standard division (SD), median and interquartile range (IQR), and for categorical variables (e.g., age groups, sex, MOT, CD4 count groups), frequency and percentages were reported. For trend analysis of categorical variables (including sex, MOT, levels of first CD4 cell counts, and age groups) over the three time periods, we used Chi-square for trend and Fisher exact tests. Kruskal–Wallis H test was used for comparing the distribution of CD4 count between the three time periods. Stata version 17 was used for epidemiological data analysis.

### Ethical consideration

This study protocol was reviewed and approved by the Research Ethics Committee of Kerman University of Medical Science (Ethics ID: IR.KMU. REC.1399. 618).

## RESULTS

Between September 1997 and December 2020, 459 HIV-positive cases have been registered in Kerman VCT. The number of registered cases in time 1 (1997-2004), 2 (2005-2012), and 3 (2013-2020) were 129, 183, and 147 cases, respectively. Of this, 105 cases with a residential address outside Kerman city and 41 homeless were excluded from the spatial analysis. Therefore, 313 cases (68.2% of all diagnosed cases) remained in the spatial analysis. However, analysis of epidemiological patterns was performed on all 459 cases.

### Epidemiological pattern

The mean (SD) age was 36.1 (9.0) years overall, 36.7 (8.1) years in men, and 34.4 (11.0) years in women. The mean age was not significantly different between the three time periods (*P* = 0.07). However, the mean age of HIV-positive males significantly increased from 34.9 years in 1997-2004 to 36.6 in 2005-2012 and 39.8 years in 2013-2020 (*P*= 0.004; Table 1).

**Table 1:** Epidemiological Characteristics of newly diagnosed cases during three period of study

| Ordinal/Nominal<br>Variables | Total (N=459)<br>n (%) | 1997-2004<br>(N=129)<br>n (%) | 2005-2012<br>(N= 183)<br>n (%) | 2013-2020<br>(N=147)<br>n (%) | P-value |
| --- | --- | --- | --- | --- | --- |
| Age Groups (Years) |  |  |  |  |  |
| ≤ 5 | 10 (2.2) | 1 (0.8) | 1 (0.5) | 8 (5.4) | 0.01 |
| 5 - 14 | 3 (0.7) | 0 | 2 (1.1) | 1 (0.7) |  |
| 15 - 24 | 24 (5.2) | 10 (7.8) | 6 (3.3) | 8 (5.4) |  |
| 25 - 34 | 157 (34.2) | 50 (38.8) | 75 (41) | 32 (21.8) |  |
| 35 - 44 | 196 (42.7) | 59 (45.7) | 70 (38.3) | 67 (45.6) |  |
| 45 ≤ | 69 (15) | 9 (7) | 29 (15.8) | 31 (21.1) |  |
| Modes of transmission |  |  |  |  |  |
| unsafe injection | 222 (48.4) | 84 (65.1) | 111 (60.7) | 27 (18.4) | <0.001 |
| sexual contact | 139 (30.3) | 15 (11.6) | 50 (27.3) | 74 (50.3) |  |
| Mother to child | 12 (2.6) | 1 (0.8) | 3 (1.6) | 8 (5.4) |  |
| Unknown | 86 (18.7) | 29 (22.5) | 19 (10.4) | 38 (25.9) |  |
| Available first CD4 count | 278 (50.6) | 28 (21.7) | 112 (61.2) | 138 (93.9) |  |
| First CD4 cell count |  |  |  |  |  |
| <200 | 116 (41.8) | 12 (42.9) | 54 (48.2) | 50 (36.2) |  |
| 200-350 | 59 (21.2) | 4 (14.3) | 20 (17.9) | 35 (25.4) |  |
| 350-500 | 37 (13.3) | 7 (25.0) | 12 (10.7) | 18 (13) | 0.23 |
| >500 | 66 (23.7) | 5 (17.8) | 26 (23.2) | 35 (25.4) |  |
| Gender |  |  |  |  | <0.001 |
| Male | 333 (72.5%) | 117 (90.7) | 140(76.5) | 76 (51.7) |  |
| Female | 126 (27.5) | 12 (9.3) | 43 (23.5) | 71 (48.3) |  |
| **Numerical Variables** | **Total (N=459)** | **1997-2004** | **2005-2012** | **2013-2020** |  |
|  | **Mean (SD)** | **(N=129)** | **(N= 183)** | **(N=147)** |  |
|  |  | **Mean (SD)** | **Mean (SD)** | **Mean (SD)** | **P-value** |
| Age Mean (SD) (Year) |  |  |  |  |  |
| Female | 34.4 (11) | 30.1 (11.3) | 35.7 (10.8) | 34.5 (11.1) | 0.37 |
| Male | 36.7 (8.1) | 34.9 (6.3) | 36.6 (8.0) | 39.8 (10.1) | 0.004 |
| Total | 36.1 (9.0) | 34.3 (7.1) | 36.4 (8.7) | 37.4 (10.8) | 0.07 |
| **CD4 (cells/ml) (Median (IQR)** | 260 (101 - 484) | 250 (78 - 438) | 248.5 (108 -480) | 281 (109 - 514) | 0.53 |

Most of the 459 cases were male (n=333, 72.5%) (Table 1). The proportion of female cases significantly increased over time, i.e., from 9.3% in 1997-2004 to 23.5% in 2005-2012 and 48.3% in 2013-2020, (*P*< 0.001). The proportion of HIV infected children under 5 years of age increased from 0.8% in 1997-2004 to 5.4% in 2013-2020 (*P* = 0.01). Unsafe injection (48.4%) and unprotected sex (30.3%) were the main MOT overall. The proportion of sexual contact as the MOT significantly increased from 11.6% in 1997-2004 to 27.3% in 2005-2012 and 50.3% in 2013-2020 (*P* < 0.001). Overall, the median (IQR) of the first CD4 count was 260 (101 - 484) cells/ml; in 41.7% of cases, the first CD4 count was under 200 cells/ml. The median of the first CD4 count or proportion of patients with the first CD4 count under 200 cells/ml did not change significantly over time.

### Spatial pattern

The NNI for newly diagnosed cases was 0.67 in 1997-2004, 0.57 in 2005-2012, and 0.53 in 2013-2020 which was clearly less than one (i.e., an indication of clustered pattern) ^14^(Figure 1). The result of the Kernel density estimation showed that overall, the distribution of newly diagnosed cases of HIV was more concentrated in the eastern parts of the city (Figure 1). The density map of residents with HIV infection had the highest density (6.01-9 cases per KM^2^) in the northern and southern edges of the city in 2005-2012.

**Figure 1:**
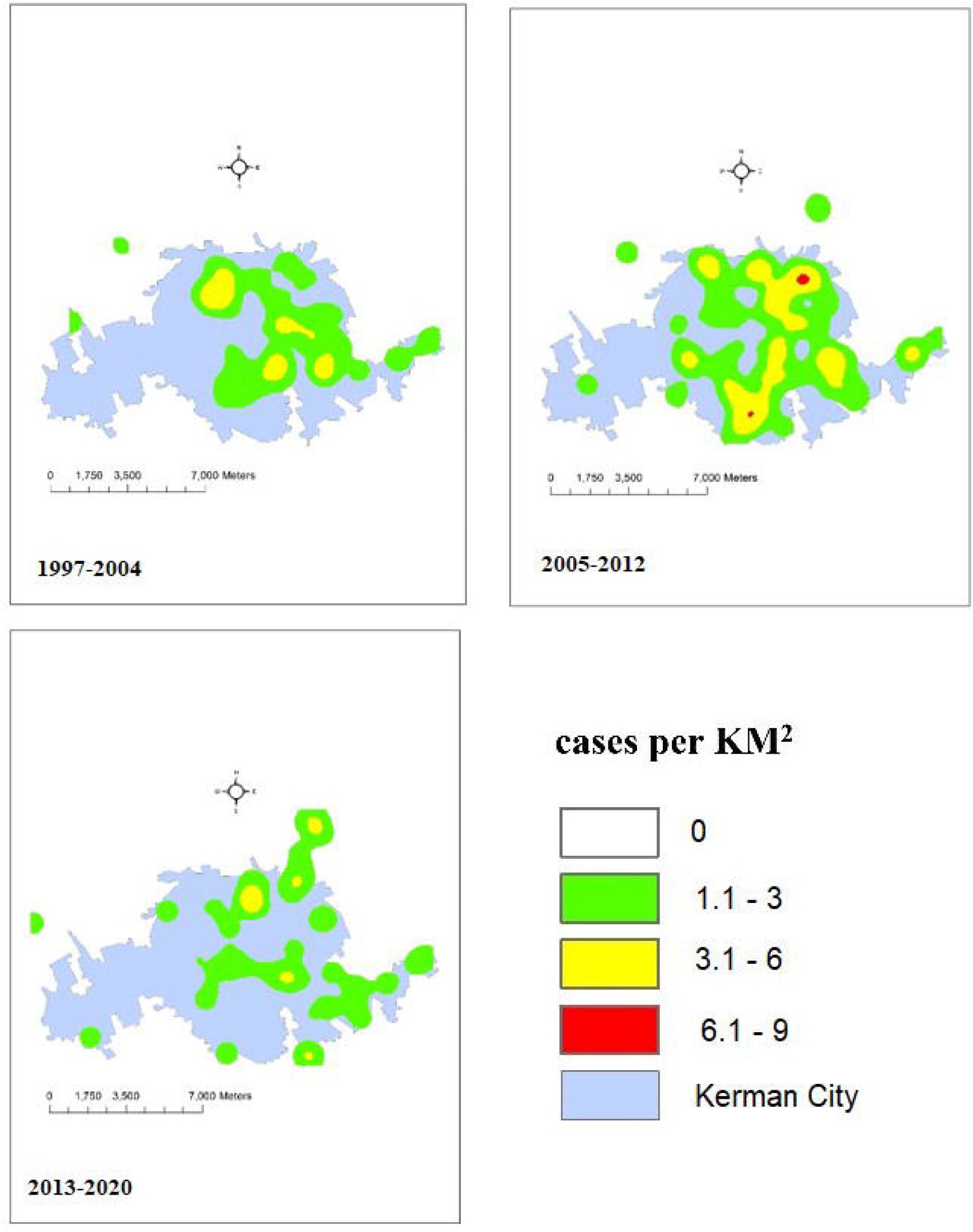
spatial distribution of people diagnosed with HIV over the study period in Kerman city

The NNI for PLWIH in 2020 was 0.63 which is suggesting a cluster pattern. HIV density map in 2020 (Figure 2) shows a high density in areas corresponding to locations of VCT, DICs, and two mobile clinics.

**Figure 2:**
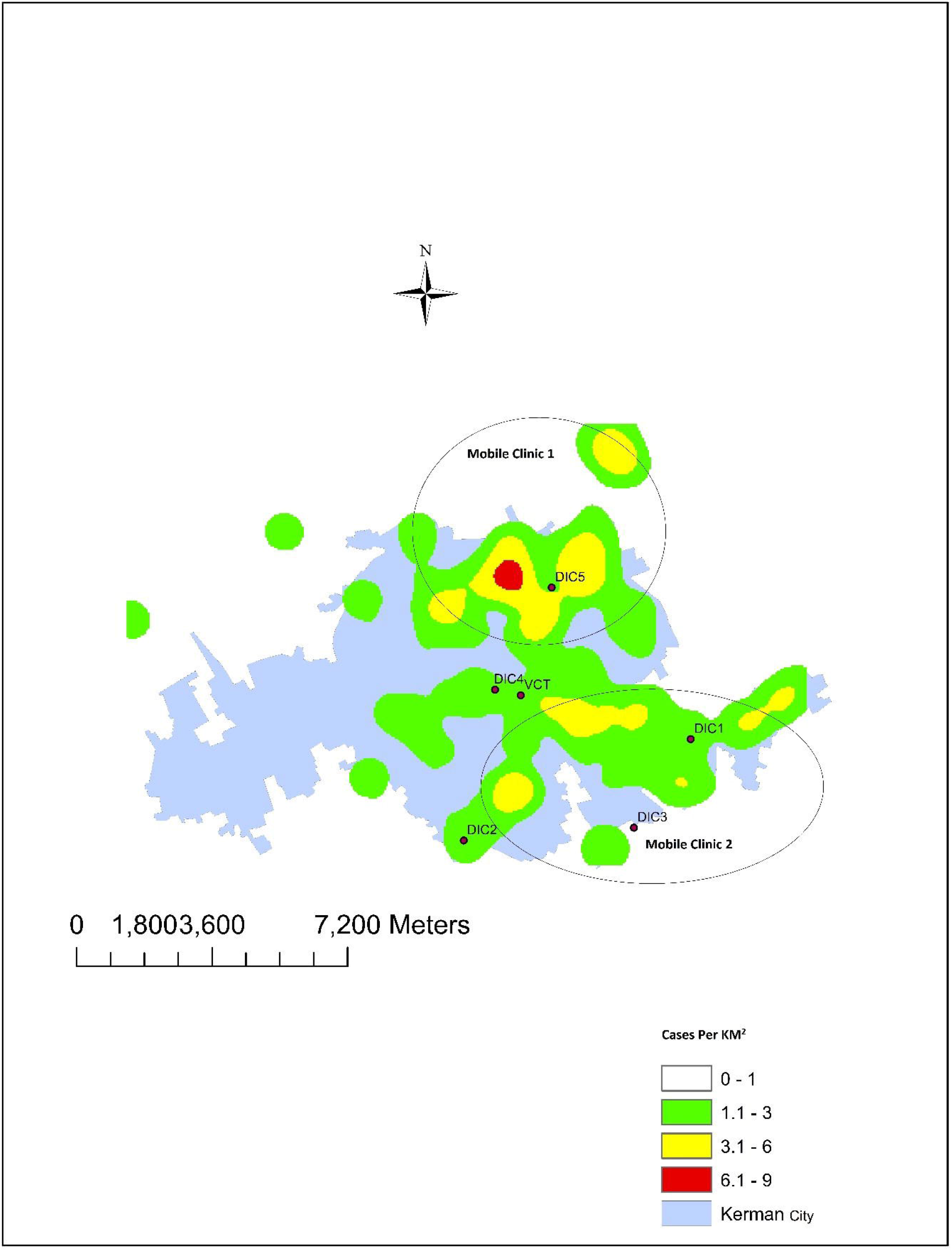
spatial distribution of people living with HIV in Kerman city in 2020

## DISCUSSION

We observed important changes in HIV epidemiology regarding gender, MOT, number of pediatric patients, and density maps over time in Kerman since 1997. The proportion of women, proportion of people infected through sexual contact, proportion of pediatric patients, and the age of diagnosed cases among male individuals increased over the time. We also found more than one in three patients had late diagnosis (CD4 count at diagnosis < 200) which has not been improved over the time. regarding the spatial analysis, we found HIV was clustered in the eastern neighborhoods of the city; density map also showed there is a shift over time with the highest density in the northern and southern edges of the city in 2005-2012. The HIV clusters in 2020 were matched with the locations of fixed and mobile related services.

The HIV is shifting from men to women and from injection to sexual routes of transmission. In this study, around three-fourth of the reported cased in the whole studied period were male, however, this proportion in the last time period was almost close to one. In total, around half of the reported cases were due to unsafe injection and less than one-third of them were due to sexual contact, however, more than half of the reported cases in the last period was due to sexual contact, and less than one-fifth of the cases were due to unsafe injection. This changing pattern from male to female and from the unsafe injection to sexual contact led to increase the number of reported cases due to mother to child (from one case at the first time period to three cases in the second time period and eight cases in the third period). This pattern is similar to the pattern observed in the national^16^ and also in some other cities in Iran, Kermanshah city (for gender) ^17^ and Yazd city (for transmission route).^18^ This changing pattern in the gender and MOT could be explained by overall change in sexual behaviors in the country over the last two decades. Having more than two clients among FSW in the last month increased from 46.6% in 2010 to 63.4% in 2015; ^5^ extramarital sex among youth girls increased from 8% in 2008 ^19^ to more than 13% in 2018; ^20^ and injection drug use among FSW increased from 14% in 2011^21^ to 25% in 2015, ^22^. However, there was no significant increase on the condom use among female sex work (condom use in the last month was 26.1% in 2010 and it was 26.3% in 2015). ^5^ Reduction of reported cases due to injection can be explained by the drug related harm reduction program in the country which are more accessible and utilized by male drug users. Methadone Maintenance Treatment (MMT) centers increased from 5,893 in 2014 to more than 8,000 centers in 2019. The number of drug users who have received MMT also increased from 476,232 individuals in 2011^16^ to more than 750,000 individuals in 2019.^23^ Moreover, shared injection among PWID decreased from 36.9% in the last month in 2010 ^1^ to 4.1% in the last three months in 2020.^3^

We also found more than three-fourth of diagnosed cased had a CD4 count ≤ 500 at diagnosed time, and more than 40% had a CD4 count <200 at diagnosis. These figures were almost similar during three time periods which shows continuous problem of late diagnosis of majority of the infected cases. In recent years, several interventions implemented in Iran to improve HIV testing. For example, HIV rapid tests have been implemented as a routine part of counseling services in health services and harm reduction settings. ^24^ HIV testing protocol also updated based on WHO guidelines that suggested health care providers to offer HIV testing as part of routine medical care to all people attending their facilities. Also, mobile HIV testing clinics were developed to increase access to HIV testing for vulnerable populations with limited access to healthcare facilities. ^25^ Due these interventions, HIV testing uptake in the last 12 months increased from 27% in 2010 to 70.4% in 2015 among FSW, ^25^ and from 24% in 2010 to 71% in 2019 among PWIDs. ^23^ Despite the overall increase in HIV testing, early detection rates have not been improved significantly. ^23^ It is not clear how many of those who tested for HIV continue to test themselves frequently as long as they are at risk or belong to a high-risk group. Repeated HIV testing recently (in 2021) added to the national HIV testing guideline, in which recommended members of high-risk groups (including; sexual partners of PLWH, PWID and their sexual partner, FSW, MSM, and prisoners) to be tested for HIV every three months. If implemented properly, it is expected that this strategy increases the chance of diagnosis HIV at early stages of infection. Proportion of key populations at risk for HIV who tested for HIV frequently also need to be monitored by surveys and surveillance efforts.

We also found the diagnosed cases were clustered in the eastern neighborhoods of the city. A study conducted in another city of Iran (Kermanshah) showed also spatial clusters of HIV cases ^17^. The higher density of HIV diagnosed cases in the eastern parts of the city can be explained by higher concentration of population, lower socioeconomic status, and higher rate of the high-risk behaviors in these neighborhoods. The density of the diagnosed case also shifted over the time with the highest density in the northern and southern edges of the city in 2005-2012. Shifting the highest density in the northern and southern edges of the city can be explained by development of a new transmission cluster in these neighborhoods. This spatial clustering of the cases could help the healthcare providers to focus on these clusters for a better case finding, identifying high risk groups and directing preventive interventions based on high-risk groups in these neighborhoods. We also found fixed and mobile health services are match closely with these clusters in 2020. There is one VCT in the city which has been established in the center of the city and it seems the location of the VCT in terms of access to all hotspots is reasonable. DICs also founded close to the hotspots and approximately cover all areas with a higher density of HIV-infected patients; the overlap between these DICs is low. DICs provide harm reduction services to the vulnerable population in place or by outreach services.

We acknowledge the limitations of our study. First, we used HIV registry data in our analysis which had missed data on some of the key variables (CD4 at diagnosis was missing in almost half, MOT was missing in one-fifth, and residential address was missing in about 10% of all cases. Although missing has been improved over time, we were not able to investigate the pattern and reasons for the missing. Second, we were not able to differentiate the heterosexual transmission from homosexual transmission routes due to the way this data were collected and reported. Finally, density map of HIV patients based on residential address may not necessarily match with the risk maps where the sexual or injection risk are taking placed.

## CONCLUSION

In this study we used HIV surveillance data from all people diagnosed with HIV in Kerman between 1997 and 2020 to assess the trend in the HIV epidemiological and spatial patterns. Our epidemiological and spatial analysis showed significant change in the HIV epidemiology regarding gender, MOT, pediatric cases, and density maps over the time. Including, increases in the proportion of women, proportion of people infected through sexual contact, pediatric cases, and the age of diagnosed cases among male individuals. This data can help to precise targeting of HIV prevention, testing, treatment and outreach programs in Kerman.

## Data Availability

ll data produced in the present study are available upon reasonable request to the authors

## Acknowledgment

We wish to acknowledge the support from the University of California, San Francisco’s International Traineeships in AIDS Prevention Studies (ITAPS), U.S. NIMH, R25MH123256. The content is solely the responsibility of the authors and does not necessarily represent the official views of the National Institutes of Health.

## Contributors

HM contributed to study design, literature review, data cleaning, data analysis, and preparation of early drafts of the paper. AM contributed to study design, analysis interpretation, critical review of the early draft of the paper. MB, contributed to study design, and preparation of early drafts of the paper. ER contributed to data extraction. SE contributed to study designs, and data analysis. HSH, managed the implementation of the study, and contributed in study design, data analysis, preparation of early drafts of the paper. All authors contributed to the writing of the paper and approved the final version.

